# BFCI at #SMM4H 2023: Integration of Machine Learning and TF-IDF for Covid-19 Tweets Analysis

**DOI:** 10.1101/2023.11.18.23297862

**Authors:** Hamada Nayel, Nsrin Ashraf, Mohamed Aldawsari

## Abstract

Extracting information from texts generated by users of social media platforms becomes a crucial task. In this paper, we describe the systems submitted to the SMM4H shared tasks 1 and 2. The aims of these two tasks are binary and multi-class classification of English tweets. We developed a machine learning-based model integrated with TF-IDF as a feature extraction approach. Four classification algorithms have been implemented namely, support vector machines, passive-aggressive classifier, multi-layer perceptron and random forest. For task 1, the passive-aggressive classifier reported f1-score of 63.7%. For task 2, multi-layer perceptron reported f1-score of 71.4%.

## Introduction

The Social Media Mining for Health Applications (SMM4H) workshop in the last couple of years has been utilized as a competitive platform for the development and evaluation of reliable Natural Language Processing (NLP) systems. The workshop aims to identify, extract, and standardize health-related information from publicly accessible user-generated content on online platforms^1^. In the last three years, Covid-19, was a global health pandemic. This pandemic attracts people to share their thoughts and opinions about their health and the extent to which they are affected by the news spreading about the disease. For the 8^th^ Session of the workshop shared task containing five tasks, the tasks contain data from Reddit and Twitter posts reporting Covid-19. Our team has participated in two tasks, task 1^2^ and task 2^3^. SMM4H 2023 task 1 – Binary Classification of English Tweets, the binary classification task involves data from automatically distinguishing tweets that self-report a Covid-19 diagnosis and data from positive test clinical diagnoses or hospitalization. SMM4H 2023 task 2 - multi-class classification, including chatter about therapies for health conditions. The task aimed at building a model easily classifies the sentiment associated with therapy into three classes (positive – neutral – negative).

## Methods

The primary aim of this work is to explore the impact of different machine learning classifiers and unigram and bBi-gram TF-IDF. All the models were implemented using *scikit-learn* environment and **NLTK**, “**re**” libraries for pre-processing. Our model comprises three phases pre-processing, feature engineering, model creation, and evaluation as shown in Figure 1.

**Figure 1.**
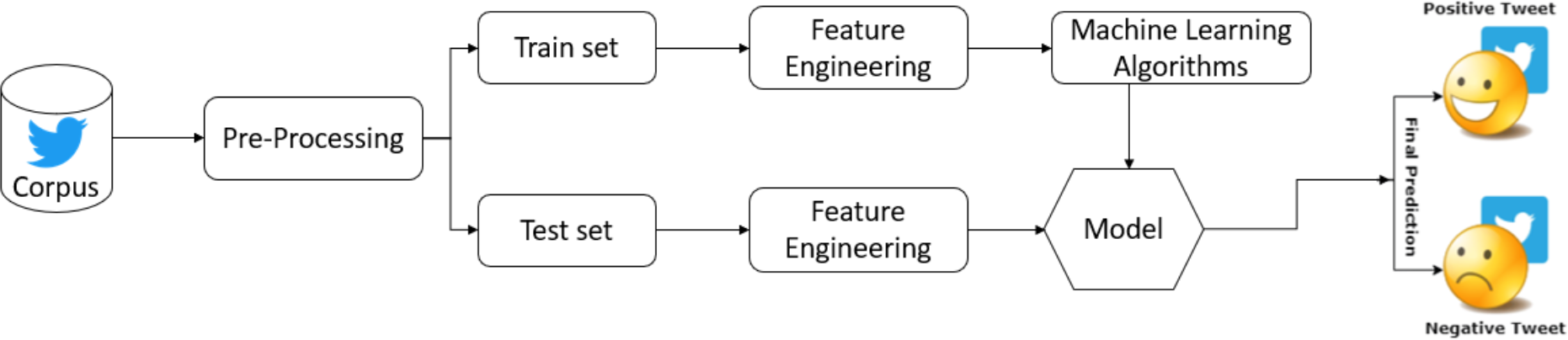
The main structure of our model.

### Pre-Processing

First, some basic data processing steps have been applied using **NLTK** and **re** packages, these steps include:

- Stopwords removal
- White special characters and non-ASCII characters removal
- White spaces and URL removal

The same preprocessing steps were applied for the two tasks.

### Features Engineering

Feature extraction is an important phase for handling data to build a classification model that is directly applied after preprocessing. In our model, Term Frequency – Inverse Document Frequency (TF-IDF) technique with a Bi-gram vector space model was applied. The model for both tasks was fine-tuned using **CV= 5, Max_DF = 1**.**0, Ngram range (1**,**2)**

### Model

In this phase, we review the methodology of the proposed models, after applying TF-IDF as a text vectorization model for text representation. Different machine-learning classification models were applied to train the data. The set of classification algorithms that have been used are listed below^4^.

### Support Vector Machine (SVM)

SVM is one of the most effective supervised machine learning algorithms used for classification. In SVM, the algorithm identifies the hyperplane that effectively distinguishes the various classes of data points. The best hyperplane is chosen based on its ability to maximize the margin between the closest data points belonging to different classes.

Regarding the hyper-parameters, the model for both tasks was fine-tuned with **linear kernel, random state =100 and scale gamma**.

### Passive Aggressive (PA)

PA is one of the most effective machine learning classifiers that solve binary classification problems, as the more it’s simple the more it’s effective. In PA, the algorithm examines the data and considers both the correct data of training samples and the aggressiveness of updates based on the magnitude of the loss. This aggression parameter allows the classifier to be aggressive in updating its parameters.

Regarding the hyper-parameters, the PA classifier applied in Task 1 was fine-tuned with **max_iter = 110, loss = ‘hinge’ and tol = ‘1e-4’**.

### Multi-Layer Perceptron (MLP)

MLP is a type of neural network structure comprising interconnected layers of neurons, designed to handle intricate tasks like pattern recognition and classification.

Regarding the hyper-parameters, MLP applied in task 2was fine-tuned with the following values of paramaters **hidden_layer =20, activation function= ‘logistic’, random_state = 42 and solver = ‘adam’**.

### Random Forest (RF)

RF is a supervised machine learning classifier in an advanced form of decision trees. RF combines multiple decision trees to make predictions based on the class with the most votes.

Regarding the hyper-parameters, RF applied in task 2 was fine-tuned with the following parameters setting **n_estimators =300 and max_depth = 5**.

## Discussion and Conclusions

In the test phase, the shared task organizers provided participants with a blind test set containing 10K tweets for task 1 and 5K tweets for task 2

### Task 1

Table 1 shows the precision, recall, and F1-score at the test phase for different classifiers. Showing that the PA classifier outperforms the SVM classifier with a 63.7% F1-score.

**Table 1.** Model Performance for Task 1 on Test Set.

| Classifier | Precision | Recall | F1-score |
| --- | --- | --- | --- |
| SVM | 0.765 | 0.540 | 0.633 |
| PA | 0.784 | 0.537 | 0.637 |

### Task 2

Table 2 shows the precision, recall, and F1-score at the test phase for different classifiers. Showing that the SVM classifier outperforms the other classifiers with a 71.4% F1-score.

**Table 2.** Model Performance for Task 2 on Test Set.

| Classifier | Precision | Recall | F1-score |
| --- | --- | --- | --- |
| MLP | 0.7141 | 0.7141 | 0.7141 |
| SVM | 0.7091 | 0.7091 | 0.7091 |
| RF | 0.6973 | 0.6973 | 0.6973 |

## Discussion and Conclusions

This paper represented a comparative study of different machine learning classifiers on #SMM4H 2023 shared tasks on tasks 1 and 2 for Health applications. The tweets have been represented as TF-IDF unigram-bigram vectors. Applying different approaches may improve the performance of the model such as Deep learning based on word embedding techniques or transformer-based models. In task 1 PA classifier achieves better performance than SVM, while in task 2 MLP achieves better performance.

## Data Availability

All data produced in the present study are available upon reasonable request to the authors

